# NC-COVID: A Time-Varying Compartmental Model for Estimating SARS-CoV-2 Infection Dynamics in North Carolina, US

**DOI:** 10.1101/2022.10.21.22281271

**Authors:** Paul L. Delamater, Rachel L. Woodul

## Abstract

Efforts to track and model SARS-CoV-2 infection dynamics in the population have been complicated by certain aspects of the transmission characteristics, which include a pre-symptomatic infectious phase as well as asymptomatic infectious individuals. Another problem is that many models focus on case count, as there has been (and is) limited data regarding infection status of members of the population, which is the most important aspect for constructing transmission models. This paper describes and explains the parameterization, calibration, and revision of the NC-COVID model, a compartmental model to estimate SARS-CoV-2 infection dynamics for the state of North Carolina, US. The model was developed early in the pandemic to provide rapid, up-to-date state-level estimates of the number of people who were currently infected, were immune from a prior infection, and remained susceptible to infection. As a post modeling exercise, we assessed the veracity of the model by comparing its output to SARS-CoV-2 viral particle concentrations detected in wastewater data and to estimates of people infected using COVID-19 deaths. The NC-COVID model was highly correlated with these independently derived estimates, suggesting that it produced accurate estimates of SARS-CoV-2 infection dynamics in North Carolina.

## INTRODUCTION

A major obstacle in efforts to mitigate and prevent SARS-CoV-2 infection and COVID-19 disease has been the presence of people with asymptomatic infections and pre-symptomatic transmission, which has (and continues to) complicate efforts to track and model infection dynamics in the population (Gao et al., 2021). The complications arise because 1) transmission cannot be traced leading to unobserved transmission of the virus and 2) unobserved infections are not captured via disease surveillance mechanisms thus requiring models for estimating the true number of infections. While confirmed case counts are available from most government agencies, these values represent only those who have been infected, accessed testing, and received a positive test result from an entity that reports their results to some centralized agency. This issue has been present throughout the pandemic, but was especially problematic in its early days when the testing infrastructure was still under development and when testing was only available to those demonstrating symptoms of COVID-19 (Perkins et al., 2020a). A recent example occurred during the Omicron wave in late 2021 and early 2022 when the use of rapid at home antigen tests increased rapidly (Rader et al., 2022), but did not require reporting (and thus was not captured by surveillance systems).

Population level epidemiological models, at a bare minimum, require estimates of the number of people who are infectious at any given time, as this value (and not the number of people who are confirmed cases) drives transmission dynamics. Unfortunately, this is not the metric captured in the COVID-19 testing and surveillance apparatus, which largely focused on people experiencing COVID-like symptoms or those with potential exposure to someone that tested positive for SARS-CoV-2 infection. Although lab confirmed infections (case counts) are a useful metric and can be used to estimate the true number of infections, there are a number of factors that affect the relationship between infections and confirmed cases including 1) the age distribution of the population because it impacts the presence of asymptomatic infections (Nikolai et al., 2020), 2) a host of aspects regarding testing such as the availability (ease of) of testing, the number of tests performed, the effectiveness of the testing approach (e.g., who gets tested), and the accuracy of the tests themselves, and 3) the number of people truly infected. While some of these factors likely remain relatively constant over time, others have varied widely over the course of the pandemic (e.g., Brown and Walensky, 2020). Essentially, the most important aspect of infection dynamic models (people infected and infectious) has been (and is) unknowable and the parameters that could be used to estimate infections from confirmed case counts have varied over time (Omori et al., 2020).

Another recurring setback for SARS-CoV-2 infection modeling efforts has been the lack of ground truth data that can be used to compare and validate modeled results (e.g., estimates of people who are infectious, remain susceptible, or have immunity via vaccination or prior infection) (Holmdahl and Buckee, 2020). This problem is especially formidable when paired with the relatively short time periods in which estimates and forecasts must be available to be useful for decision-making purposes, as well as the evolving nature of the pandemic and scientific understanding of SARS-CoV-2 transmission. This issue was exceptionally problematic during early efforts to plan and implement COVID-related mitigation efforts, as there was considerable uncertainty regarding many aspects of the virus and its characteristics (Holmdahl and Buckee, 2020).

In the early stages of the pandemic, we developed the NC-COVID model, a compartmental model to estimate SARS-CoV-2 infection dynamics for the state of North Carolina, US. The motivation for building the NC-COVID model was to provide rapid, up-to-date state-level estimates of the number of people who were currently infected, were immune from a prior infection, and remained susceptible to SARS-CoV-2 infection. The model results were updated daily and made freely available on the website https://www.nc-covid.org. Because the NC-COVID model was developed and deployed early in the COVID-19 pandemic, we made numerous revisions and improvements, especially as more (and better) information and data became available.

In this paper, we describe the NC-COVID model and explain its parameterization, calibration, and revision over time. We focus on the initial and final forms of the model. As a post modeling exercise, we compare the output to SARS-CoV-2 viral particle concentrations detected in wastewater data and to estimates of people infected using COVID-19 deaths, offering an additional assessment of the veracity of the model.

## MODEL DESCRIPTION

### Overview

The overall goal of the modeling effort was to use publicly available data to produce up-to-date estimates of the number of people newly infected with SARS-CoV-2, as well as the number currently infectious, susceptible, and immune, as these values drive short term infection dynamics in a population and are not readily available from surveillance systems. While we eventually used the model to make short-term forecasts, the focus of the effort was to estimate the “current” conditions based on the most up-to-date data; we updated our model each day that new data was released, which was daily for much of the modeling period.

### Structure

We used a deterministic compartmental model to estimate state-level SARS-CoV-2 infection dynamics (Anderson and May, 1991). In the initial stage of development and implementation (prior to the availability of vaccines), the model included compartments for susceptible (S), exposed (E), infectious (I), removed (R, recovered from infection and immune), and dead (D). Because the initial modeling effort was planned to take place only over short period (e.g., 6-9 months), we did not account for demographic changes in the population due to immigration, emigration, births, or natural deaths in the model.

Once the COVID-19 vaccine became available, we added compartments for people who received a single dose of the two dose series vaccine and became immune (V_1I_), people who received a single dose of the two dose series vaccine and remained susceptible (V_1S_), and people who received the second dose of the two dose series and became immune (only those who did not become immune after the first dose) or received the single dose vaccine and became immune (V_2I_).

### Infection Parameters

A compartmental model requires parameters to estimate the flow of people between the various compartments over time. In an SEIR model, these include the effective contact rate *β*, rate of loss of latency *δ*, and the recovery rate *γ*. The *δ* parameter is simply the inverse of the incubation period (length of time that a person is infected but not yet infectious), while the *γ* parameter is the inverse of the infectious period (length of time that a person is infected and infectious, i.e., able to spread the infection). The Infection Fatality Rate (*IFR*) describes the proportion of people who die from the infection.

During model development, inspection of the statewide daily lab confirmed cases suggested that SARS-CoV-2 transmission dynamics were altered following three distinct time points corresponding to statewide nonpharmaceutical interventions (NPIs). Hence, to account for various NPIs and behavioral changes that affected transmission, we allowed *R*_0_ to vary over time in the SEIR model. The incubation period and infectious period were not allowed to vary over time thus the *δ* and *γ* parameters did not change. As such, the effect of varying *R*_0_ over time was essentially to allow the *β* parameter to vary (*β* = *R*_0_ * *γ*). We divided the study period into distinct temporal periods using breakpoints (where noticeable deviations from a prior trend occurred), which were identified by visually examining the lab confirmed case count data along with a list of potential actions having the potential to affect transmission of SARS-CoV-2 on a state level (e.g., when the statewide mask mandate went into effect). To implement this approach in the SEIR model framework, we solved the differential equations in a piecewise fashion (separately for each temporal period), using the number of people in each compartment at the end of one period as the starting state for the next temporal period (Hou et al., 2020). Generally, we attempted to limit the number of breakpoints to the minimum number necessary to capture major shifts in transmission. Moreover, it is important to note that this approach did not require that *R*_0_ change at a breakpoint, but allowed it to if supported by the observed data.

### Vaccination

When vaccine administration began in December 2020, we added compartments to the SEIR model for vaccinated individuals as described above. We used observed vaccination data to determine the number of people who received their initial dose of an mRNA vaccine (*V*_*obs,1*_) and a single dose of the Johnson and Johnson vaccine (*V*_*obs,S*_); we assumed that the vaccinated individuals were drawn proportionally from the Susceptible and Recovered populations (people with an active SARS-CoV-2 infection are not recommended to receive the vaccine). We used a temporal lag to account for the time between receiving the vaccine and developing immunity and effectiveness multipliers to assign immune status to those who developed immunity after the first dose of an mRNA vaccine series (*ρ*_*1*_), after the second dose of an mRNA vaccine series (*ρ*_*2*_), and after a single dose of the Johnson and Johnson vaccine (*ρ*_*S*_). We used a temporal lag to account for the recommended time between doses of the mRNA vaccine series (*L*); we assumed some people who received the first dose of an mRNA vaccine would not receive the second dose (Lost to Follow up, LTF). The compartments and flows among them for the entire model are detailed in Figure 1.

**Figure 1:**
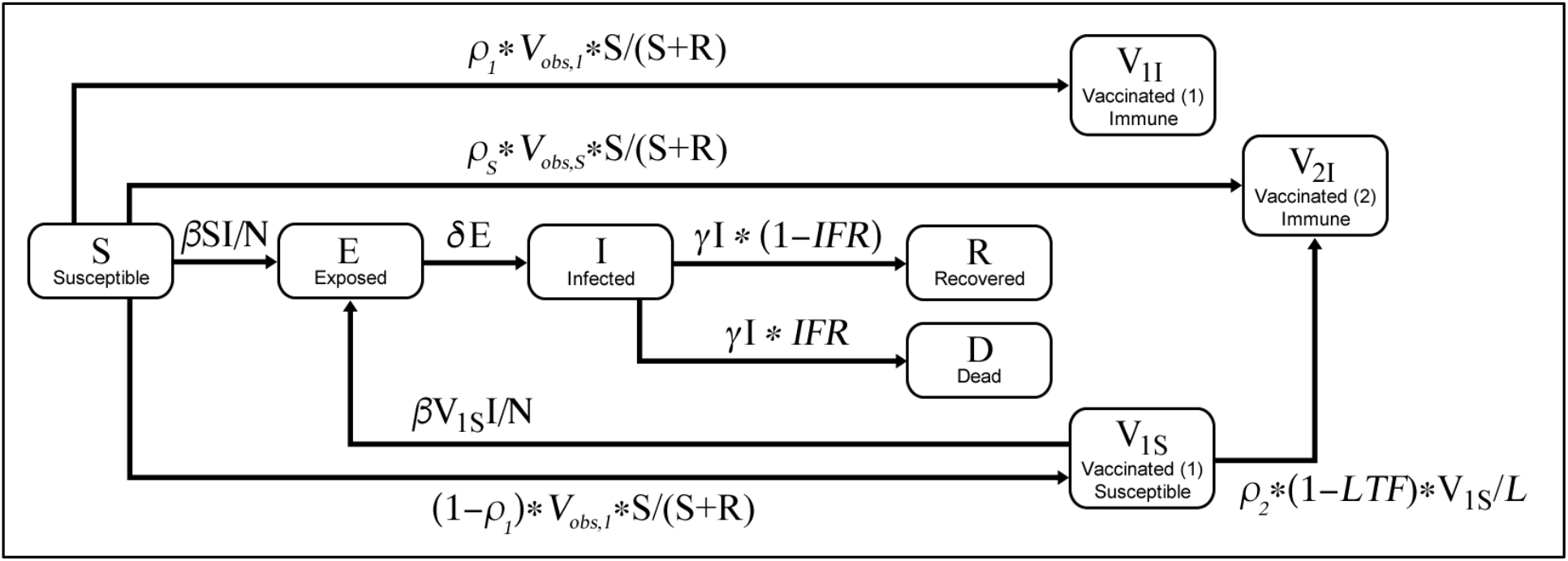
Compartments and equations showing the flow of people among compartments in the final version of the SARS-CoV-2 model for North Carolina.

### Data

All data used in later versions of the model, which included the daily number of reported cases, tests reported, and people vaccinated, were drawn from the North Carolina COVID-19 Dashboard (North Carolina Department of Health and Human Services, NCDHHS, n.d.). Earlier versions of the model used cases and testing data from NCDHHS provided by WRAL (WRAL, 2020).

## MODEL CALIBRATION

### Overview

To calibrate the model, we allowed numerous model parameters to vary given the high level of uncertainty (especially early in the pandemic). To account for this, we implemented a two-stage calibration approach that included an initial and tuning phase. We evaluated the goodness of fit of the model by estimating daily lab confirmed infections from the compartmental model output and comparing them to the observed data. The initialization and tuning process was implemented during major revisions of the model. When inserting a new temporal period, we estimated the *R*_0_ value for the two most recent time periods using only the tuning phase of the calibration process. Daily model updates were restricted to estimating the *R*_0_ value of the most recent temporal period.

### Model Calibration and Updates

We calibrated the parameters in our model using a two-stage approach. In the initial stage, we used Latin Hypercube Sampling (McKay et al., 1979) to draw random values for the parameters from a uniform distribution with relatively large ranges for each model run. We did this for 300,000 model runs to account for the high levels of uncertainty that characterized the early part of the pandemic. From the results of the initial stage, for each parameter value, we calculated the mean and standard deviation using the values from the 5,000 most accurate runs. In the tuning phase, we ran the model an additional 30,000 times while randomly drawing parameter values from a normal distribution based on the mean and standard deviation values calculated in the initial phase. We then chose the single set of parameter values that produced the best fit to the observed data for the final model. This process was reimplemented when a major revision was made to the model.

When incorporating a new temporal period, we performed a simplified calibration process, only updating the R_0_ values for the two most recent temporal periods. The other parameter values were unchanged from the most recent implementation of the full calibration process. Daily model updates were limited to updating the R_0_ value of the most recent temporal period.

### Lab Confirmed Infections and Testing

To compare the results of our model to observed data required estimating the number of lab confirmed cases that would result from the number of people who were infected, as this was one of the only metrics available (and thus usable for calibration purposes). However, the number of lab confirmed cases is highly dependent on the nature of the testing infrastructure, and a person infected with SARS-CoV-2 might not be identified as a lab confirmed case for several reasons including 1) not getting tested because they are asymptomatic, 2) not getting tested because testing was not available to them at the time, and 3) getting tested after a potential exposure, but prior to detectability (Reese et al., 2021).

During the initial phase of model development and implementation (in May 2020), while the COVID-19 testing program was ramping up, we estimated the percent of all new infections confirmed via testing as a parameter that linearly increased based on point estimates calculated for the first date of data (March 13^th^, 2020) and the most recent date of observed data at the time the model was run (two parameters). A later version of the model estimated the percent of infections that were lab confirmed as a linear rescaling function of the number of tests performed each day (parameters based on endpoints of 1,000 and 40,000 daily tests performed). We calculated the 7-day floating average of the daily number of tests reported given the noisiness of the daily data. We lagged the 7-day average of daily tests by four days prior to July 7^th^, 2020 to account for the longer delay in processing and reporting test results very early in the pandemic (no lag afterwards). To calculate lab confirmed cases from the compartmental model output, we multiplied the estimated daily number of new people infected (those moving into the Exposed compartment) by the estimated percent of infections confirmed via testing (based on the estimated number of tests that day).

### Assessing Goodness of Fit

We assessed the model’s goodness of fit using the root mean squared error (RMSE) of the daily estimated lab confirmed cases compared with observed data from NCDHHS. We used the 7-day floating average of the observed data given the high amount of day-to-day noise in the raw data. The observed case data were lagged by seven days to account for the delay between when a person was infected and when they would be reported as a positive case.

### Initial Date and Conditions

Although the North Carolina Health and Human Services reported the first lab-confirmed SARS-CoV-2 infection on March 3, 2020 (NCDHHS, 2020), SARS-CoV-2 was likely circulating prior to this initial positive test result. Preliminary efforts to initialize the model with a low number of infections the week prior to this date did not produce plausible results suggesting there was a far higher number of people infected before the first case was reported. As such, we initialized our model on February 24, 2020 with 200 people in the infectious compartment and 400 people in the exposed compartment. Later work showing that the SARS-CoV-2 virus may have been circulating in the US population well before late January 2020 supported this approach (Basavaraju et al., 2021).

### Temporal Periods

In the initial version of the model, we included four time periods with breakpoints assigned at the beginning of the statewide Stay-At-Home order (March 30, 2020), the beginning of Phase 1 of the “Staying Ahead of the Curve” reopening plan (May 8, 2020), and the beginning of Phase 2 reopening plan (May 22, 2020). As we continued to update the model and transmission conditions changed (e.g., lifting of NPIs or during the phased reopening plan), we added additional periods. The dates of statewide actions were gathered from NCDHHS (NCDHHS, n.d.). In an attempt to distinguish between true changes and small deviations that may have been influenced by data anomalies, we waited approximately 1-2 weeks after a noticeable deviation between the observed data and the modeled estimates or a visible deviation in the trend of the daily case data (e.g., a pronounced peak or valley) to add an additional breakpoint. The final set of temporal periods with a short description of the event (or events) associated with the beginning of each are presented in Table 1.

**Table 1:** Temporal periods with start (breakpoint date) and description.

| Period | Start date | Description |
| --- | --- | --- |
| 1 | February 24, 2020 | Start date |
| 2 | March 30, 2020 | Beginning of Stay-at-Home order |
| 3 | May 8, 2020 | Beginning of Phase 1 of reopening plan |
| 4 | May 22, 2020 | Beginning of Phase 2 of reopening plan |
| 5 | July 6, 2020 | Beginning of statewide mask mandate, post-July 4<sup>th</sup> holiday |
| 6 | September 4, 2020 | Beginning of Phase 2.5 of reopening plan |
| 7 | November 23, 2020 | Thanksgiving holiday |
| 8 | January 4, 2021 | Post-New Year Holiday |
| 9 | March 4, 2021 | Modified Stay-at-Home order lifted, further easing of restrictions |
| 10 | June 21, 2021 | Introduction of Delta variant |
| 11 | August 4, 2021 | Decrease in transmission |
| 12 | September 8, 2021 | Further Decrease in transmission |

### Calibration Results

The first version of the NC-COVID model was run for the period of February 24, 2020 to June 11, 2020 (the most up-to-date data at the time the model was run). Model fit was evaluated from March 13, 2020 (the first date with the number of tests reported) to June 11, 2020. The set of parameters from the initial model calibration are presented in Table 2, including the parameter ranges used for the initial calibration phase (initial parameter ranges were drawn from Lauer et al., 2020; Ma et al., 2020; Perkins et al., 2020b), the mean and standard deviation of the parameters used in tuning calibration phase, and the resulting values of the best fit model. The RMSE of the daily lab confirmed case data in the initial version of the model was 25.61 (cases per day).

**Table 2:** Parameter values from the initial model including the range used in the initial stage, the mean and standard deviation used in the tuning stage, and the final values from the best fit model.

| Parameter | Unit | Initial |  | Tuning |  | Final |
| --- | --- | --- | --- | --- | --- | --- |
|  |  | Low | High | Mean | Std Dev | Tuned |
| Incubation Period | Days | 4 | 8 | 5.77 | 1.11 | 5.85 |
| Infectious Period | Days | 5 | 12 | 7.42 | 1.63 | 6.09 |
| Infections Lab Confirmed, 3/13/20 | Percent | 4 | 10 | 7.87 | 1.55 | 7.44 |
| Infections Lab Confirmed, 6/11/20 | Percent | 10 | 25 | 17.33 | 4.28 | 20.38 |
| $R_0$ (Period 1) | Unitless | 2 | 3 | 2.69 | 0.23 | 2.67 |
| $R_0$ (Period 2) | Unitless | 1 | 2 | 1.28 | 0.16 | 1.13 |
| $R_0$ (Period 3) | Unitless | 1 | 2 | 1.26 | 0.17 | 1.17 |
| $R_0$ (Period 4) | Unitless | 1 | 2 | 1.36 | 0.18 | 1.27 |

The final “full” calibration of the model was performed on October 9, 2020 using data from March 13, 2020 (the first date with the number of tests reported) to October 8, 2020 (the most up-to-date data at the time). The results are presented in Table 3. In this version of the model, the RMSE of the daily case estimates was 98.45 (cases per day).

**Table 3:** Parameter values from the final full calibration of model including the range used in the initial stage, the mean and standard deviation used in the tuning stage, and the final values from the best fit model.

| Parameter | Unit | Initial |  | Tuning |  | Final |
| --- | --- | --- | --- | --- | --- | --- |
|  |  | Low | High | Mean | Std Dev | Tuned |
| Incubation Period | Days | 5 | 7 | 5.86 | 0.54 | 5.99 |
| Infectious Period | Days | 5 | 7 | 5.80 | 0.52 | 6.05 |
| Infections Lab Confirmed, 1,000 tests | Percent | 4 | 10 | 7.31 | 1.68 | 10.89 |
| Infections Lab Confirmed, 40,000 tests | Percent | 30 | 40 | 35.03 | 2.88 | 37.13 |
| $R_0$ (Period 1) | Unitless | 2.5 | 2.9 | 2.74 | 0.11 | 2.73 |
| $R_0$ (Period 2) | Unitless | 1 | 1.25 | 1.15 | 0.07 | 1.17 |
| $R_0$ (Period 3) | Unitless | 1 | 1.3 | 1.16 | 0.09 | 1.25 |
| $R_0$ (Period 4) | Unitless | 1.1 | 1.4 | 1.25 | 0.07 | 1.21 |
| $R_0$ (Period 5) | Unitless | 0.8 | 1.2 | 0.98 | 0.04 | 0.99 |
| $R_0$ (Period 6) | Unitless | 1 | 1.4 | 1.16 | 0.10 | 1.04 |

### Vaccination Parameters

We lagged the observed data of people who received a first dose of an mRNA vaccine by 14 days to account for time delay from vaccine administration to immunity; we estimated that 60% of susceptible people would gain immunity after the first dose (*ρ*_*1*_) (Britton et al., 2021). We assumed that the people receiving this first dose were drawn proportionally from the Susceptible and Recovered population at that time. Those who were already immune via prior infection (in the Recovered compartment) were not moved from that compartment (regardless of whether they were scheduled to gain immunity via vaccination), and those who were vaccinated but did not gain immunity were not moved from their respective compartment. We used a waiting period of 24.5 days from the date of receiving the first dose of an mRNA vaccine to receipt of a second dose. This represents the average of the recommended time between doses for the Pfizer (28) and Moderna (21) vaccine series. We assumed that 95% of people would receive their second dose of an mRNA vaccine, meaning 5% would be lost to follow-up (Kriss et al., 2021). We included an additional 14-day lag after receipt of the second dose prior to developing immunity. We estimated that 80% of those who received both doses of an mRNA vaccine would develop immunity (*ρ*_*2*_) (Baden et al., 2021; Dagan et al., 2021; Polack et al., 2020). Since 60% of people had already gained immunity after the first dose in our model, this meant 50% of those who did not acquire immunity after the first dose acquired it after the second dose. For those that received the single dose COVID vaccine, we assumed 80% would gain immunity after 14 days (Sadoff et al., 2021).

### Final Results

The final version of the model was run on October 15, 2021 using data from March 3, 2020 to October 14, 2021. The R0 values for the first five time periods are found in Table 3; the *R*_0_ values for time periods 7 to 12 were 1.19, 1.44, 1.01, 1.61, 3.76, 2.94, 2.33. The final version of the model resulted in an overall RMSE of 287.2 (cases per day) based on daily lab-confirmed cases. Observed lab confirmed cases and the modeled results for the entire study period are presented in Figure 2.

**Figure 2:**
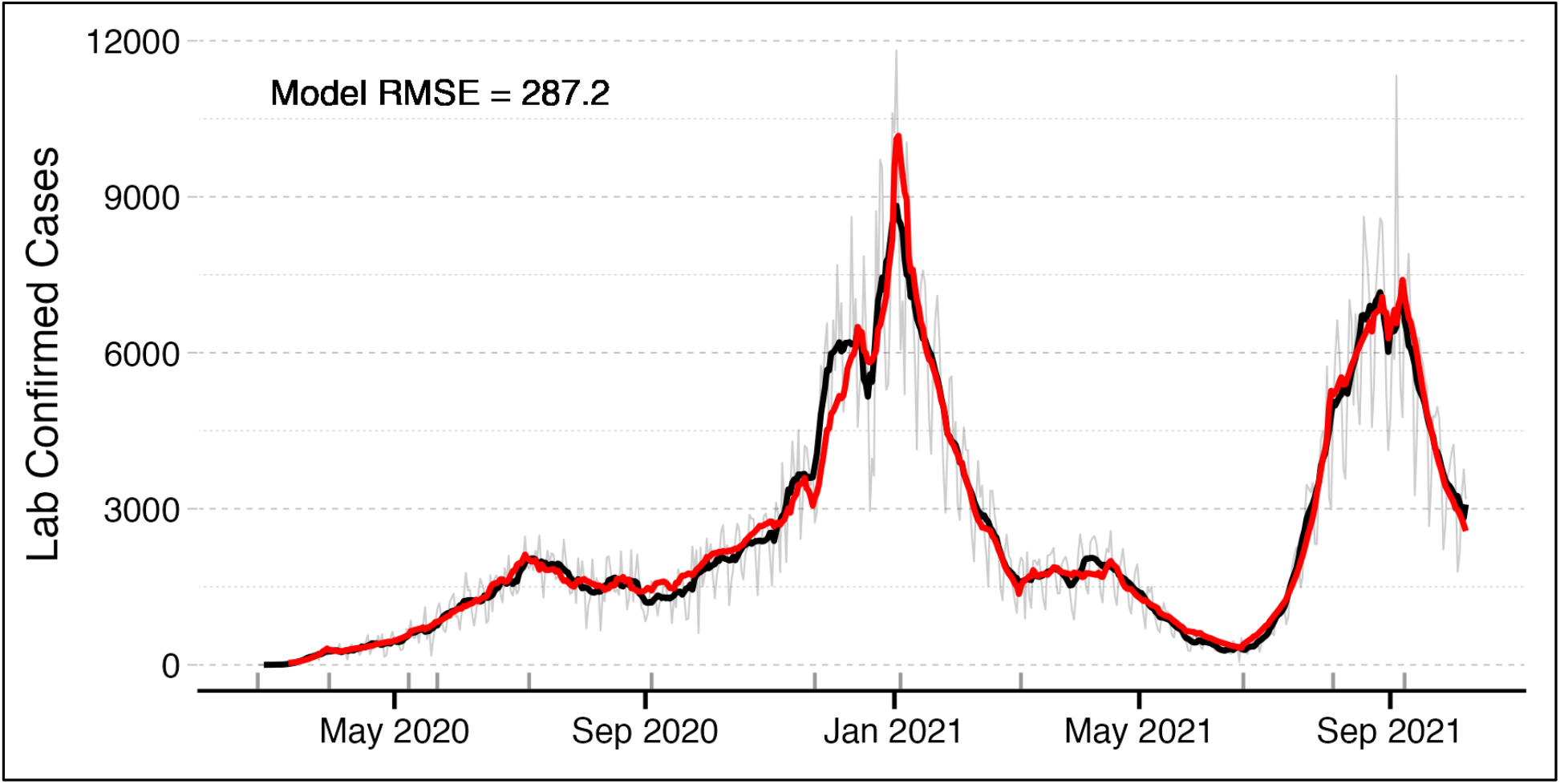
Observed lab confirmed COVID-19 cases (grey line), 7-day rolling average of lab confirmed cases (black line), and modeled results (red line) in North Carolina. The temporal breakpoints (Table 1) are shown using grey lines on the X axis.

The estimated percent of SARS-CoV-2 infections that were lab confirmed over the study period is presented in Figure 3. Given that this is a linear rescaling of the observed testing data, the temporal pattern is quite similar in nature to that data. The overall magnitude of the values appears feasible, as the NC-COVID model estimated that just over 10% of all SARS-CoV-2 infections were being confirmed via testing in the early stages of the pandemic and just over 60% were being confirmed when the number of infections reached its (pre-Omicron) peak (when roughly 60,000 to 80,000 test results were being reported daily).

**Figure 3:**
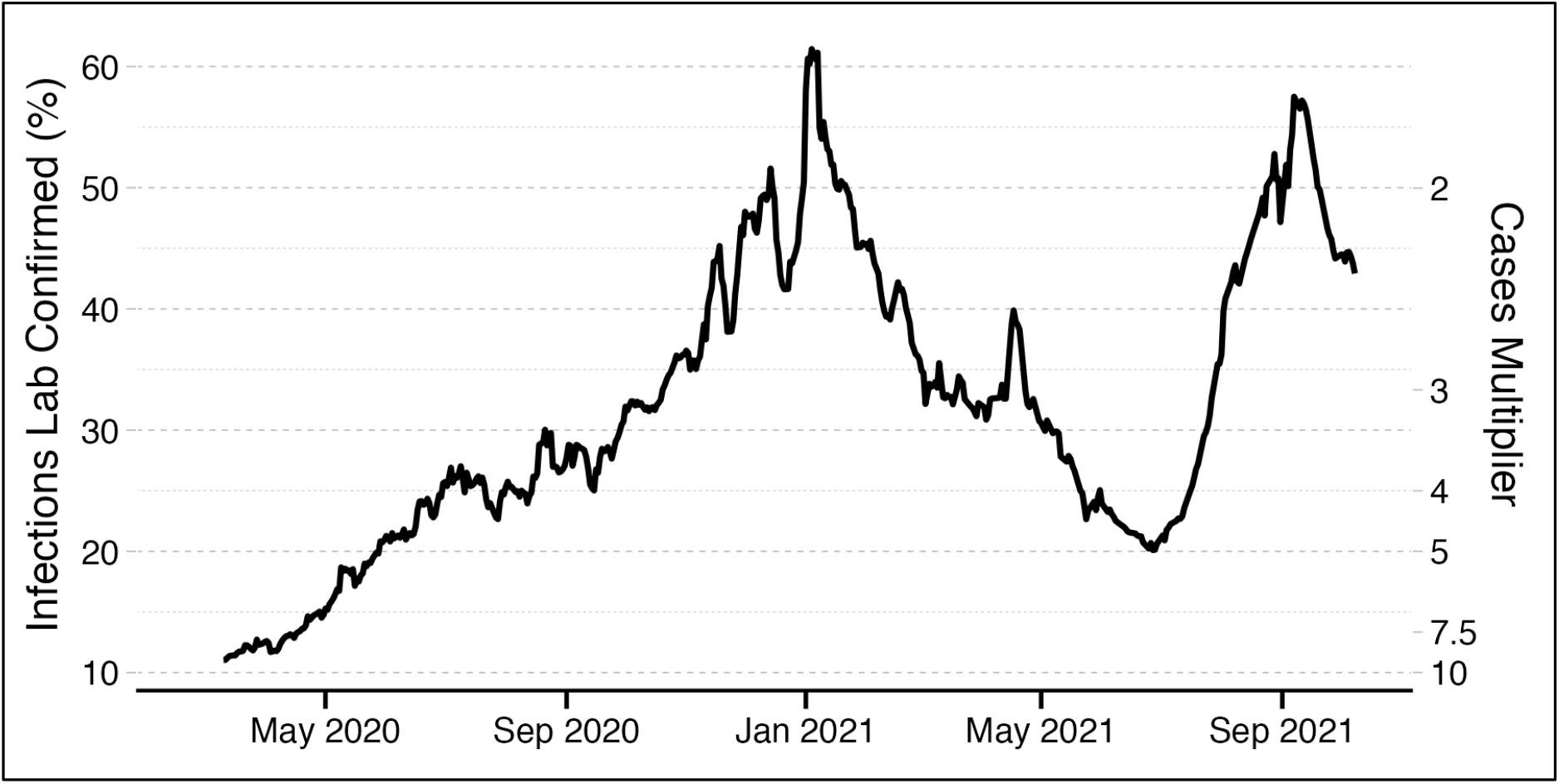
Estimated percent of true SAR-CoV-2 infections confirmed via testing in North Carolina. For reference, the cases multiplier to calculate infections is provided on the right side Y axis (the inverse of percent confirmed).

A simplified version of the compartment distribution of NC’s population in the NC-COVID model is shown in Figure 4. For display purposes, people who were vaccinated but remain susceptible (V_1S_) are included with greater susceptible (S) population, the people who are exposed (E) and those infectious (I) are combined, those who gained immunity after a first dose of the mRNA vaccines (V_1I_) are grouped with those who gained it after the second mRNA dose (V_2I_) and those who gained it after the first dose of the Johnson and Johnson vaccine (also included in V_2I_). Finally, those who died (D) are included with those who recovered from infection (R). When viewed in the context of the entire state population and over a roughly 20-month period, the peaks and valleys in Figure 2 are quite muted, and the people gaining immunity via SARS-CoV-2 infection steadily increased over time. A similar observation can be made with regards to those gaining immunity via vaccination; despite the ebbs and flows in the number of people vaccinated since the first vaccinations in December of 2021, the cumulative number of people with immunity increased at a relatively steady pace. In the last run of the NC-COVID model (which was completed prior to the surge in infections of the SARS-CoV-2 Omicron variant), there was (roughly) a similar proportion of NC’s population in each of the three simplified compartments (Susceptible, Immune from vaccination, and Recovered from infection).

**Figure 4:**
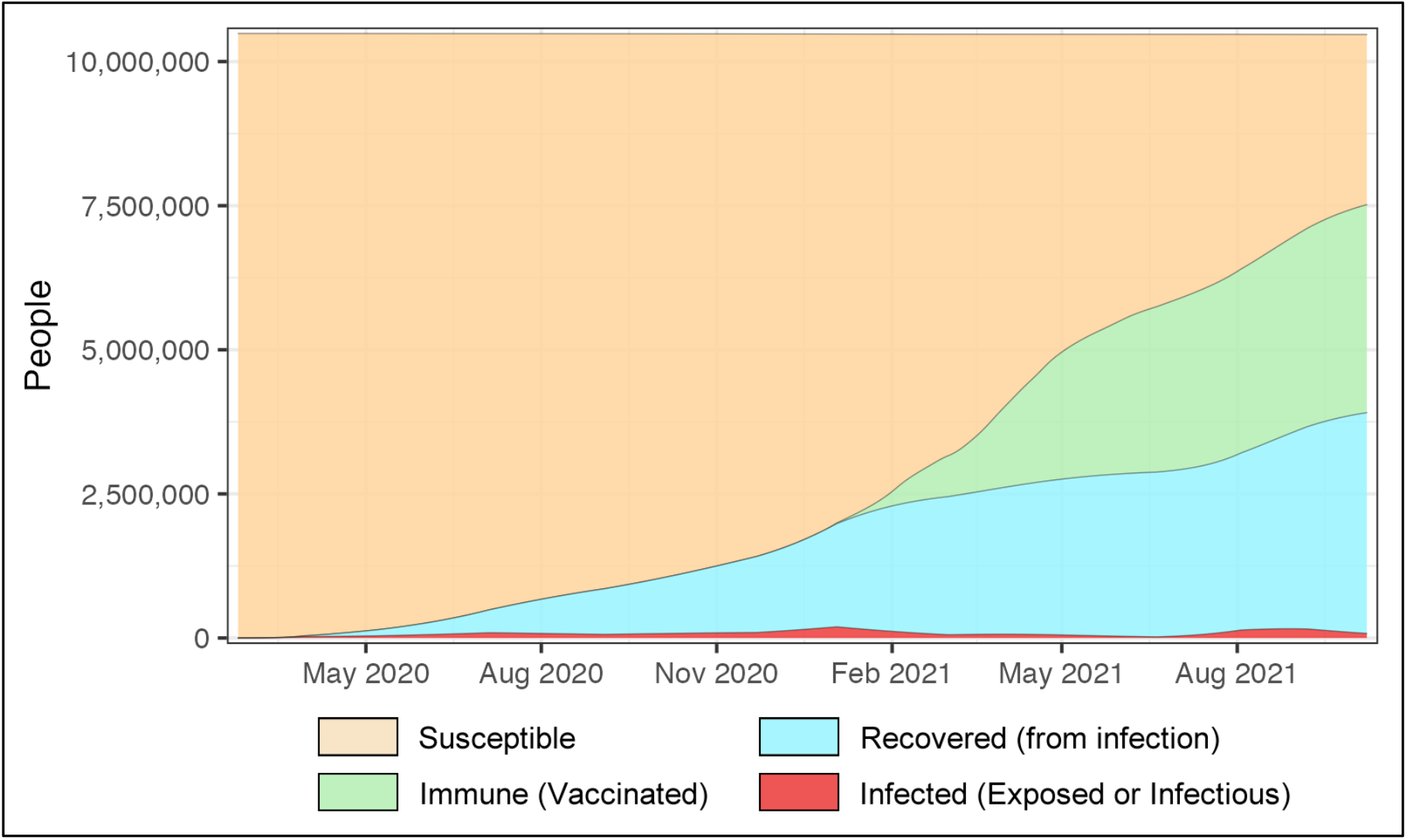
Summary of people in NC-COVID model compartments over time. Susceptible includes those who had not been infected and those who had been vaccinated but had not developed immunity. Immune includes those who gained immunity via vaccination (but not infection). Recovered includes those who gained immunity via infection (but not vaccination). Infected includes people in the Exposed compartment (not yet infectious) as well as the Infectious compartment.

## MODEL VALIDATION

During the development and dissemination period of the NC-COVID model, our focus was on providing rapid, up-to-date estimates of infections in NC (we updated the model every day that new data were released by NCDHHS). Hence, at the time, we did not conduct model validation using any outside data. Given an opportunity to look back on the model, we conducted a post hoc model validation exercise using two external data sources, wastewater surveillance data and reported COVID-19 deaths.

### Wastewater Surveillance Data

We compared the model’s estimate of the current number people infected to estimates derived from wastewater surveillance data. Wastewater surveillance is a well-established method for detection and monitoring of waterborne and enteric pathogens (CDC, 2019a, 2019b; Sims and Kasprzyk-Hordern, 2020). RNA fragments from SARS-CoV-2 are reliably shed in human feces, both before the infected individual becomes symptomatic and in the absence of symptom development. As such, infectious people who were not identified via testing would be represented in wastewater surveillance if they lived within a sewershed being monitored. This *post hoc* analysis of the model’s performance provides additional layer of validation that was not available during the model development process.

Wastewater data were gathered from the North Carolina Wastewater Monitoring Network, part of the Centers for Disease Control and Prevention’s (CDC) National Wastewater Surveillance System (NWSS). This program samples wastewater from 25 sewersheds across the state, representing a mix of large, medium, and small municipalities (North Carolina Wastewater Monitoring Network, 2022) and containing roughly 22% of the state’s population (exact value has changed over time as sewersheds were added to the network). We used the viral gene copies per person metric, which is a normalized value that accounts for varying levels of viral genes in the water, the wastewater flow, and the number of people in the sewershed. The data were reported twice weekly over the study period.

We calculated the statewide value of the SARS-CoV-2 viral gene copies per person using a weighted average of the sewershed-level data (using sewershed population as the weights). We used a linear interpolation to convert the data to a daily representation (from the twice weekly observations) to match the model output, and then calculated the 7-day moving average to smooth temporal fluctuations. The data were evaluated over a period of January 3, 2021 (the first available wastewater data) to October 15, 2021 (the final available model data).

### COVID-19 Deaths Data

We compared our modeled estimates of people newly infected to estimates derived from reported COVID-19 deaths. Despite being subject to potential overcounts and undercounts, COVID-19 death data has been used for this purpose in prior analyses (Flaxman et al., 2020; McCulloh et al., 2020; Phipps et al., 2020). We adopt the general approach of Flaxman et al. to “backcast” the number of infections that would have generated the observed number of deaths. We simplified their approach using point estimates for the parameters (rather than drawing from a distribution) using the following equation: I_t-lag_ = D_t_ / *IFR* where the number of deaths that occur on any day (D_t_) are assumed to be generated by the people who were newly infected on an earlier date (I_t-lag_). Deaths are divided by the infection fatality rate (*IFR*) to estimate infections.

We used an *IFR* of 0.065%, which was calculated by averaging the values reported from the CDC (2022) (0.063%) and a meta-analysis (Meyerowitz-Katz and Merone, 2020) (0.068%). Estimates of the lag time from infection to death varies across sources, but generally are reported to be 20-25 days (CDC, 2021a; Irons and Raftery, 2021; Verity et al., 2020). We evaluated each integer value over this range and report the best fit with the NC-COVID model result.

### Validation Methodology

We used Pearson and Spearman correlation to assess the relationship between the daily number of people currently infected from the model and the SARS-CoV-2 viral gene copies per person as well as the relationship between people newly infected (entering the Exposed compartment of the NC-COVID model) and the estimates of new infections from the observed death data.

### Validation Results

The number of people infected estimate from the NC-COVID model and the SARS-CoV-2 viral gene copies per person data are plotted over time in Figure 5(A); we used a linear rescaling factor of 17.5 to align the two Y axes. Figure 5(B) contains a scatterplot with histograms of the two variables. The Pearson R was equal to 0.92, p < 0.001 (Spearman R = 0.96, p < 0.001), signaling an extremely strong relationship between estimated people infected and SARS-CoV-2 virus detected in the wastewater. The temporal plot shows a somewhat large deviation in the wastewater data around September, 2021. Because the wastewater data covers less than 25% of NC’s population, the deviation could have been due to a surge of infections occurring specifically in the reporting sewersheds.

**Figure 5:**
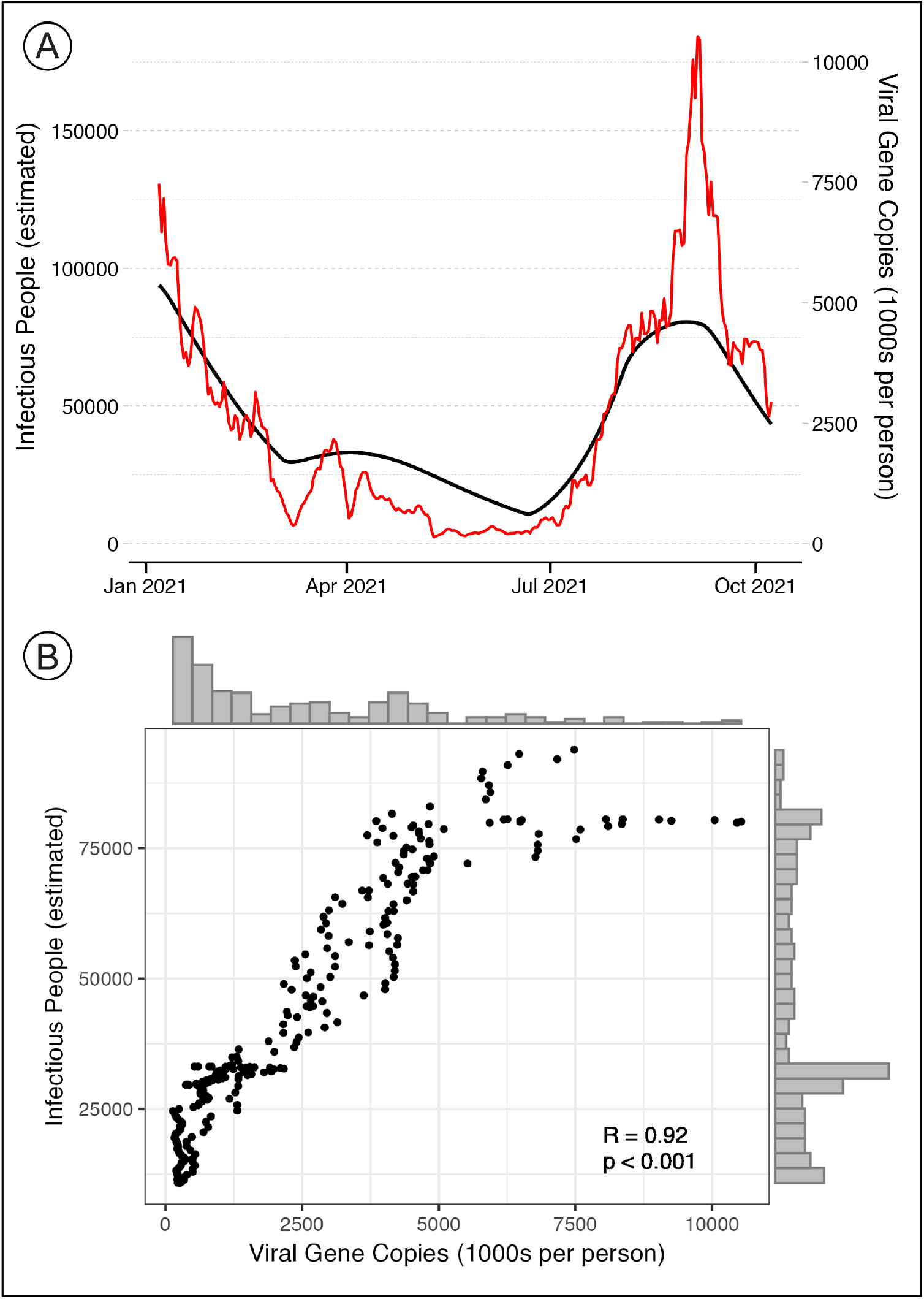
Relationship between the number of infectious people from the NC-COVID model and the viral gene copies data from the wastewater. In (A), both variables are plotted over time with the infection people plotted in black and the wastewater data plotted in red. (B) contains a scatterplot with histograms for each variable and the results of the Pearson correlation analysis (R = 0.92, p < 0.01).

The results of the comparison between new daily infections from the NC-COVID model and the backcasted estimates from observed death data are presented in Figure 6. The observed deaths were lagged by 21 days from the date of death for the estimate; however, the fit was only slightly weaker using other lag values (Supplemental Table S1). Given a lag of 21 days, the evaluation period was from February 24, 2020 to September 17, 2021. There was an extremely high level of agreement among the two estimates with Pearson R = 0.92, p < 0.001 (Spearman R = 0.89). The infection estimates from the death data are generally less than the estimates from the NC-COVID model. One explanation for this outcome is that NC had a lower IFR than the general estimate. Another notable aspect of Figure 6(B) is that the lag period appears to shift early in 2021; this may be due to improvements in COVID-19 treatment or the effects of vaccination, ultimately leading to a longer average period between SARS-CoV-2 infection and death.

**Figure 6:**
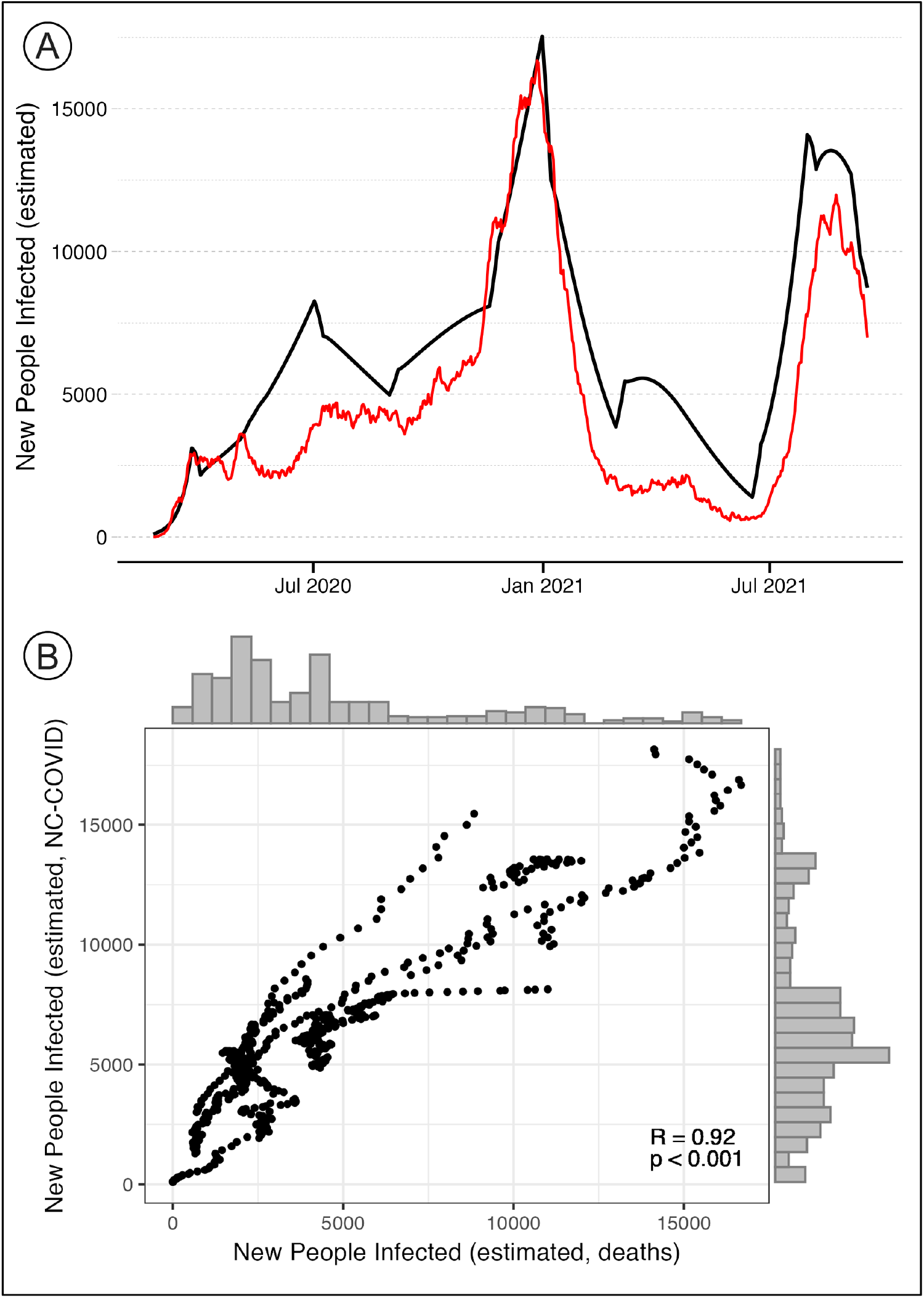
Relationship between the number of new people infected from the NC-COVID model and based on observed death data. In (A), both variables are plotted over time with the NC-COVID model estimates plotted in black and the estimates based on death data plotted in red. (B) contains a scatterplot with histograms for each variable and the results of the Pearson correlation analysis (R = 0.92, p < 0.01).

## DISCUSSION

Decision-making in the early stages of the COVID-19 pandemic was characterized by uncertainty due to the novelty of the virus and the evolving nature of its scientific and medical understanding (Paulik et al., 2020), as well as by the widespread circulation of mis- and disinformation (Sauer et al., 2021; Zarocostas, 2020). One difficult aspect for both individuals and decisionmakers to navigate seemed to be how to assess the risk of infections for various activities given the incomplete, uncertain, changing, and in some cases erroneous information. It was particularly difficult to understand the level of community prevalence and transmission occurring at this stage due a testing program that was still largely under development. A common misunderstanding at that time was that “reported cases” did not represent all people who had been infected with SARS-CoV-2 (Raubenheimer, 2020). In early to mid 2020, the CDC estimated that there were 10 people infected for every reported case (Kniesner and Sullivan, 2020). Although this may have been somewhat common knowledge at the time, estimates of the infection status of the population (e.g., susceptible, infected but not infectious, infectious, and recovered from infection and immune) were not readily available.

One of the main motivators behind the development and deployment of the NC-COVID model was to rapidly estimate the SARS-CoV-2 infection status of the NC population, and to make the information freely available to any interested parties via the nc-covid.org website. While we did not keep record of every example of use of our model estimates and website, we do know they were used (at least as a reference) by public school districts, small businesses, city and regional planners, university administrators, health care professionals, and local and state government officials. The NC-COVID model and website were also featured in more than 25 news pieces (including articles and television spots) between July, 2020 and August 2021 (https://nc-covid.org/press.html).

The presence of pre-symptomatic and asymptomatic transmission adds complexity and difficulty to population-level SARS-Cov-2 infection dynamics models. Early in the pandemic, when it was important to provide information rapidly, we developed a relatively straightforward model to accomplish this. In this work, we explain its parameterization, calibration, and revision over time in an effort to shed light on not only the importance of the model itself, but also on the process of quickly developing a model and disseminating its results early in a pandemic when finding useful, trustable information can be troublesome. While we knew at the time of modeling that the NC-COVID model results fit the observed case data quite well, the results of our post-modeling analysis demonstrate that it is highly correlated with two independent approaches to modeling the number of people who were infected thus corroborating its accuracy.

## Data Availability

All data produced in the present study are available upon reasonable request to the authors

https://www.nc-covid.org

## Notes

### Competing Interest Statement

The authors have declared no competing interest.

### Funding Statement

Funding for this project was provided by The National Institute of Allergy and Infectious Diseases (NIAID), award number 1K01AI151197-01 (PI: P Delamater).

