## Supplemental Information for "NC-COVID: A Time-Varying Compartmental Model for Estimating SARS-CoV-2 Infection Dynamics in North Carolina, US"

| LAG | PEARSON | SPEARMAN |
|---|---|---|
| 20 | 0.916 | 0.889 |
| 21 | 0.916 | 0.891 |
| 22 | 0.916 | 0.892 |
| 23 | 0.915 | 0.892 |
| 24 | 0.914 | 0.892 |
| 25 | 0.911 | 0.892 |

**Supplemental Table S1:** Pearson and Spearman R correlation values for different lags (days) from date of death for new infection estimates (compared to NC-COVID estimate). All p-values were $< 0.001$.
